# Elevated adiponectin and altered metabolic signaling in asymptomatic malaria: a community-based study of Nigerian children

**DOI:** 10.1101/2025.11.04.25339530

**Authors:** Bose Etaniamhe Orimadegun, Oluyemisi J. Adeolu, Kofoworola I. Adediran, Adebola E. Orimadegun

**Author notes:** Corresponding Author: (AEO). Senior author (Adebola E. Orimadegun).

## Abstract

Asymptomatic *Plasmodium falciparum* infection is common among school-aged children in endemic settings, yet its metabolic consequences are poorly defined. We investigated whether subclinical malaria is associated with altered adipocytokine profiles in Nigerian children and whether effects vary with parasite density. In a community-based cross-sectional survey in Ibadan, Nigeria, we enrolled 317 primary-school children and ascertained malaria by rapid test and light microscopy. Plasma adiponectin, leptin, and resistin were quantified using validated enzyme immunoassays, and the leptin-to-adiponectin ratio was derived. Multivariable linear models with clustering by school evaluated associations between asymptomatic malaria and each biomarker, adjusting for age, sex, height-for-age and body-mass-index-for-age z-scores, socioeconomic status, and hemoglobin. Sensitivity analyses restricted exposure to microscopy-confirmed infection and to participants with C-reactive protein ≤3 mg/L. Among malaria-positive children, we examined correlations between parasite density and adipocytokines with false discovery rate control.

Of 317 children, 102 (32.2%) had asymptomatic parasitemia. Compared with uninfected peers, infected children had higher adiponectin (geometric mean 45.2 vs 32.1 μg/mL) and a lower leptin-to-adiponectin ratio. In adjusted analyses, asymptomatic malaria was associated with +42% adiponectin (95% CI 18 to 71) and −31% leptin-to-adiponectin ratio (95% CI, −49 to −7); leptin and resistin were not independently associated with infection status. Among malaria-positive children, higher parasite density correlated with higher adiponectin (Spearman 0.31; 95% CI, 0.12 to 0.47), while correlations with other markers were not significant. Findings were consistent in sensitivity analyses, including after restricting to low C-reactive-protein samples.

Asymptomatic malaria in childhood is characterized by elevated adiponectin and a reduced leptin-to-adiponectin ratio, indicating altered metabolic signaling during subclinical infection. The dose–response with parasite density suggests that ostensibly silent infections are metabolically active. These results suggest considering metabolic factors in child-focused malaria control strategies and motivate tests of whether treating asymptomatic infection normalizes metabolic signals and improves longer-term health.

## Introduction

Asymptomatic malaria, detectable *Plasmodium* parasitemia without fever or other clinical symptoms, remains pervasive in sub-Saharan Africa and undermines elimination efforts by sustaining a silent human reservoir of infection [1–3]. In high-transmission settings such as Nigeria, subclinical infection is common in school-age populations and may impose a persistent, low-grade inflammatory burden despite the absence of overt illness [1,2,4–6]. Emerging evidence indicates that asymptomatic parasitemia is not immunologically inert and can intersect with developmental trajectories that matter for growth, cognition, and metabolic health in childhood [2,4–6].

A converging body of work links chronic or recurrent immune activation to altered metabolic control, immunometabolism, where inflammatory cues reshape endocrine mediators of energy balance and, reciprocally, adipose-derived hormones modulate immunity [7]. Within this interface, adipocytokines (adiponectin, leptin, and resistin) function as endocrine integrators of energy homeostasis and inflammatory signaling [8]. Leptin regulates appetite and supports pro-inflammatory immune responses [9,10], adiponectin enhances insulin sensitivity and generally exerts anti-inflammatory effects [11,12], and resistin has been implicated in inflammatory and insulin-resistance pathways [13]. Infections can perturb these adipocytokines in ways that influence host defense and tissue protection, contributing to heterogeneity across clinical states [8,14].

Prior work on malaria and adipocytokines has largely focused on symptomatic disease and adult cohorts, yielding patterns that may not generalize to the asymptomatic pediatric state [2,6]. Given the high prevalence of subclinical infection among children in West Africa and its potential metabolic consequences [1,2,4–6], clarifying how adipocytokines behave during asymptomatic parasitemia is essential to determine whether this “silent” infection is metabolically active.

Accordingly, we investigated adipocytokine profiles in Nigerian schoolchildren with and without asymptomatic malaria. Our objectives were to: (1) compare circulating adiponectin, leptin, and resistin between malaria-positive and malaria-negative children; (2) examine relationships between adipocytokines and parasite density; (3) evaluate links with nutritional status; and (4) test whether any differences persist after adjusting for age, sex, socioeconomic status, and growth indicators. Here, we characterize adipocytokine profiles in Nigerian schoolchildren with and without asymptomatic malaria and examine whether associations vary with parasite density.

## Methods

### Ethics Statement

The study was approved by the Oyo State Ministry of Health Research Ethics Committee (Approval AD13/479/016c; 2024), Ibadan, Nigeria. Written informed consent was obtained from parents/guardians and assent from children aged ≥7 years. Data were de-identified and securely stored. Reporting follows the STROBE checklist for cross-sectional studies [15].

### Study design and setting

We conducted a cross-sectional analysis of baseline (pre-intervention) data from a school-based community survey of primary-school children in Akinyele Local Government Area (LGA), Ibadan, Nigeria. Data and specimens were collected March–June 2024. Field activities and biospecimen handling were coordinated at the Institute of Child Health (ICH), College of Medicine, University of Ibadan.

### Participants and sampling

Children were recruited from 20 public primary schools within approximately 10–15 km of the ICH. Eligibility at screening included attendance on survey day, typical primary-school age, and absence of severe acute illness. Asymptomatic status at enrollment was confirmed by afebrile axillary temperature and a brief symptom screen negative for acute illness. Written parental consent and child assent were obtained prior to study procedures.

For this analysis, we included all children with complete malaria screening (light microscopy) and plasma measurements of adiponectin, leptin, and resistin; this yielded 317 children (215 malaria-negative; 102 malaria-positive). Because recruitment was school-based, subsequent models used school as the clustering unit for variance estimation.

### Definitions of exposure and outcomes

Malaria status at baseline was defined as malaria-positive if microscopy detected *Plasmodium* parasites; malaria-negative required both tests to be negative. A sensitivity analysis restricted exposure to microscopy-confirmed infection. Primary and secondary outcomes were plasma adiponectin (μg/mL), leptin (ng/mL), resistin (ng/mL), and the leptin-to-adiponectin (L/A) ratio. Among malaria-positive children, we further examined associations between parasite density (parasites/μL) and adipocytokines. For inflammation-restricted sensitivity analyses, we used C-reactive protein (CRP) ≤3 mg/L to reduce potential confounding by acute inflammatory processes (threshold consistent with CDC/AHA guidance separating low vs higher background inflammation [19]).

### Data collection and measurements

Height and weight were measured with standardized procedures; height-for-age (HAZ) and BMI-for-age (BAZ) z-scores were computed with WHO AnthroPlus using the 2007 WHO growth reference for 5–19 years [16,17]. Axillary temperature and a brief symptom screen were obtained at the time of blood draw. Capillary blood was tested by Giemsa-stained blood film microscopy following standard WHO guidance [18]. Parasite density (parasites/μL) was recorded for microscopy-positive samples.

### Blood sample collection and laboratory assays

Collection and processing. Venous blood (5 mL) was collected into EDTA tubes and transported at 4–8 °C. Plasma was separated within 4 h by centrifugation for 10 min at 3,000 × g (2–8 °C), aliquoted, and stored at −80 °C until analysis. A maximum of one freeze–thaw cycle was permitted.

Assays and quality control. Plasma adiponectin was measured using the Human Adiponectin (ADP) ELISA (Melsin Medical Co., Changchun, China; range 3.125–100 μg/mL; intra-assay CV <10%; inter-assay CV <15%). Leptin was measured using the Human Leptin ELISA (Melsin; CAT. NO: EKHU-0715; range 0.5–16 ng/mL; intra-assay CV <10%; inter-assay CV <15%). Resistin was measured using the Human Resistin ELISA (Melsin; CAT. NO: EKHU-0845; range 1.5–48 ng/mL; minimum detectable dose <0.1 ng/mL; intra-assay CV <10%; inter-assay CV <15%). CRP was measured using a high-sensitivity ELISA (R&D Systems). Complete blood counts were performed on a Sysmex KX-21N (Kobe, Japan) for a subset of participants.

All assays were run in duplicate by personnel blinded to malaria status. The mean of duplicates was used for analysis; samples with duplicate CV >15% were re-assayed. Units are reported as μg/mL (adiponectin) and ng/mL (leptin, resistin) throughout; any kit documentation originally in mg/L was converted once using 1 mg/L = 1 μg/mL for adiponectin.

### Study size and sample-size determination

This study was powered around plasma adiponectin, analyzed on the log scale to compare children with asymptomatic malaria versus uninfected peers. At design stage we specified the smallest biologically meaningful difference as a ∼30% contrast in geometric means between groups. Anticipating approximately ratio 2:1 group sizes, for malaria-negative to malaria-positive, we set two-sided α = 0.05 and 80% power and targeted ∼320 children, allowing for losses from unusable samples/data checks. Cluster-robust standard errors (by school) were prespecified to accommodate residual within-school correlation [20,21]. The achieved analytic sample (n = 317; allocation ≈2:1) met these criteria. Secondary endpoints (leptin, resistin, L/A) and interaction tests were not separately powered and are interpreted as supportive/exploratory.

### Statistical analysis

Continuous variables were assessed for distributional assumptions using histograms and Q–Q plots, complemented by Shapiro–Wilk tests. Because adipocytokines were right-skewed, values were log-transformed before parametric analyses. Descriptive results are presented as geometric means with 95% confidence intervals; in regression, effects are expressed as percent differences derived from exponentiated coefficients. Baseline comparisons used χ² tests for categorical variables and Mann–Whitney U tests for continuous variables, with standardized mean differences (SMDs) summarizing effect magnitude.

Primary analyses examined associations between malaria status and each log-adipocytokine using multivariable linear regression with cluster-robust (Huber–White) standard errors [20,21], clustering on school. Covariate adjustment proceeded hierarchically: Model 1 included age and sex; Model 2 added height-for-age and body-mass-index-for-age z-scores (HAZ, BAZ); Model 3 further adjusted for socioeconomic status and hemoglobin. Among malaria-positive children, relations between parasite density and adipocytokines were evaluated with Spearman’s rank correlation; 95% CIs were obtained from 1,000 bootstrap resamples [23]. Multiplicity was controlled using the Benjamini–Hochberg false discovery rate within prespecified test families (e.g., adipocytokines; parasite-density correlations) [22]; both raw and FDR-adjusted p-values are reported. Missing data was modest: regression models used complete-case analysis and correlations used pairwise deletion. All tests were two-sided with α = 0.05. Analyses were conducted in Stata/BE 18.0 (Stata Corp, College Station, TX) [24].

## Results

### Participant characteristics

We analyzed 317 children with complete adipocytokine measurements (215 malaria-negative; 102 malaria-positive). Baseline characteristics are summarized in Table 1. The median age was 7.2 years (IQR 4.1–9.8), and 52.4% (n=166) were female. Children with asymptomatic malaria were slightly older than uninfected peers (median 7.8 vs 6.9 years; p=0.045). Distributions of sex, nutritional status indicators, and socioeconomic characteristics were broadly similar by malaria status. Full per-variable denominators and test types are provided in S1 Table.

**Table 1:**
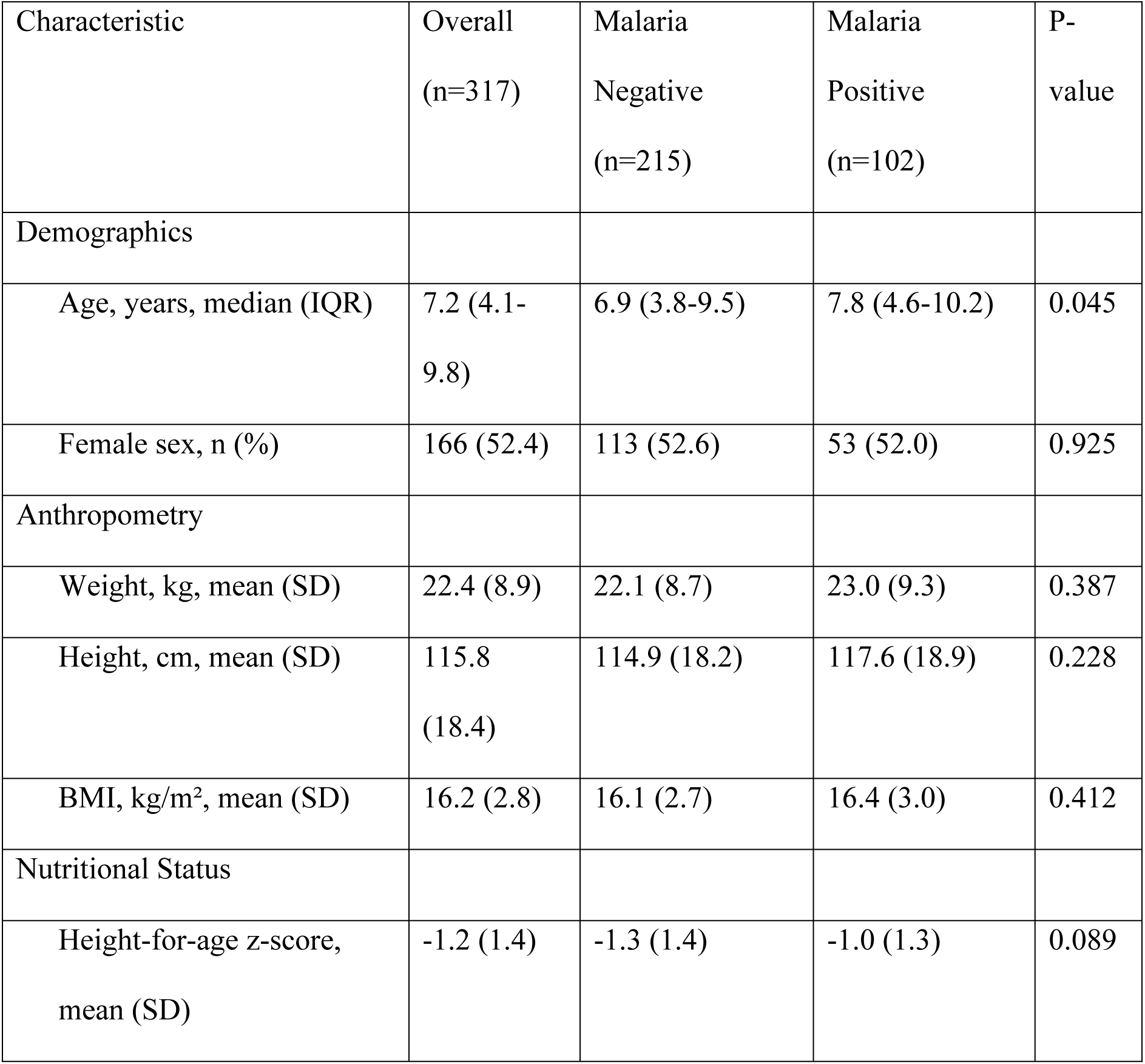
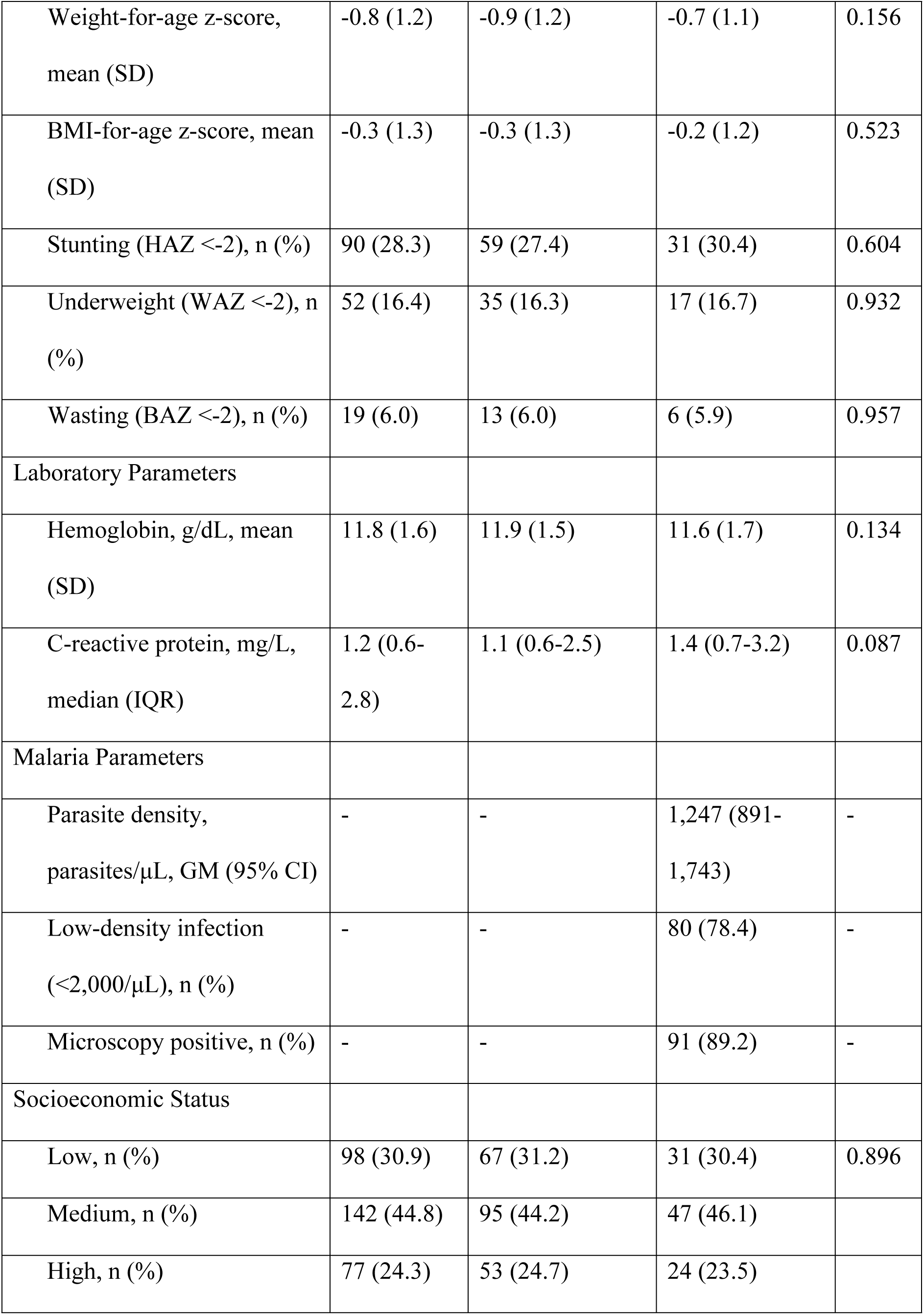
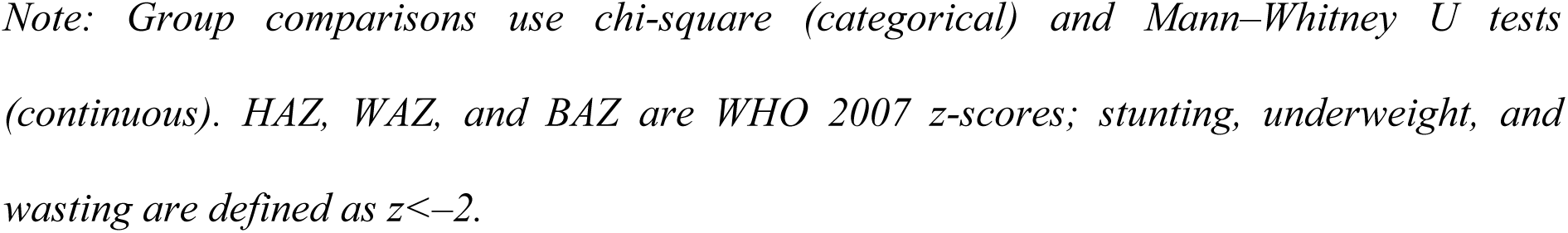
Baseline characteristics of study participants by malaria status.

| Characteristic | Overall<br>(n=317) | Malaria<br>Negative<br>(n=215) | Malaria<br>Positive<br>(n=102) | P-value |
| --- | --- | --- | --- | --- |
| Demographics |  |  |  |  |
| Age, years, median (IQR) | 7.2 (4.1-9.8) | 6.9 (3.8-9.5) | 7.8 (4.6-10.2) | 0.045 |
| Female sex, n (%) | 166 (52.4) | 113 (52.6) | 53 (52.0) | 0.925 |
| Anthropometry |  |  |  |  |
| Weight, kg, mean (SD) | 22.4 (8.9) | 22.1 (8.7) | 23.0 (9.3) | 0.387 |
| Height, cm, mean (SD) | 115.8<br>(18.4) | 114.9 (18.2) | 117.6 (18.9) | 0.228 |
| BMI, kg/m<sup>2</sup>, mean (SD) | 16.2 (2.8) | 16.1 (2.7) | 16.4 (3.0) | 0.412 |
| Nutritional Status |  |  |  |  |
| Height-for-age z-score, mean (SD) | -1.2 (1.4) | -1.3 (1.4) | -1.0 (1.3) | 0.089 |
| Weight-for-age z-score, mean (SD) | -0.8 (1.2) | -0.9 (1.2) | -0.7 (1.1) | 0.156 |
| BMI-for-age z-score, mean (SD) | -0.3 (1.3) | -0.3 (1.3) | -0.2 (1.2) | 0.523 |
| Stunting (HAZ <-2), n (%) | 90 (28.3) | 59 (27.4) | 31 (30.4) | 0.604 |
| Underweight (WAZ <-2), n (%) | 52 (16.4) | 35 (16.3) | 17 (16.7) | 0.932 |
| Wasting (BAZ <-2), n (%) | 19 (6.0) | 13 (6.0) | 6 (5.9) | 0.957 |
| Laboratory Parameters |  |  |  |  |
| Hemoglobin, g/dL, mean (SD) | 11.8 (1.6) | 11.9 (1.5) | 11.6 (1.7) | 0.134 |
| C-reactive protein, mg/L, median (IQR) | 1.2 (0.6-2.8) | 1.1 (0.6-2.5) | 1.4 (0.7-3.2) | 0.087 |
| Malaria Parameters |  |  |  |  |
| Parasite density, parasites/μL, GM (95% CI) | - | - | 1,247 (891-1,743) | - |
| Low-density infection (<2,000/μL), n (%) | - | - | 80 (78.4) | - |
| Microscopy positive, n (%) | - | - | 91 (89.2) | - |
| Socioeconomic Status |  |  |  |  |
| Low, n (%) | 98 (30.9) | 67 (31.2) | 31 (30.4) | 0.896 |
| Medium, n (%) | 142 (44.8) | 95 (44.2) | 47 (46.1) |  |
| High, n (%) | 77 (24.3) | 53 (24.7) | 24 (23.5) |  |

Note: Group comparisons use chi-square (categorical) and Mann–Whitney U tests (continuous). HAZ, WAZ, and BAZ are WHO 2007 z-scores; stunting, underweight, and wasting are defined as $z < -2$ .

### Adipocytokine concentrations by malaria status

Adipocytokine profiles differed by malaria status (Fig 1, Table 2). Malaria-positive children had higher plasma adiponectin than uninfected controls (geometric mean 45.2 μg/mL [95% CI 39.8–51.3] vs 32.1 μg/mL [95% CI 29.4–35.1]; p<0.001), with a large standardised mean difference (SMD 0.89; 95% CI 0.61–1.17). The leptin-to-adiponectin ratio (L/A) was lower in malaria-positive children (geometric mean 0.062 [95% CI 0.051–0.075] vs 0.072 [95% CI 0.064–0.081]; p=0.009).

**Fig 1.**
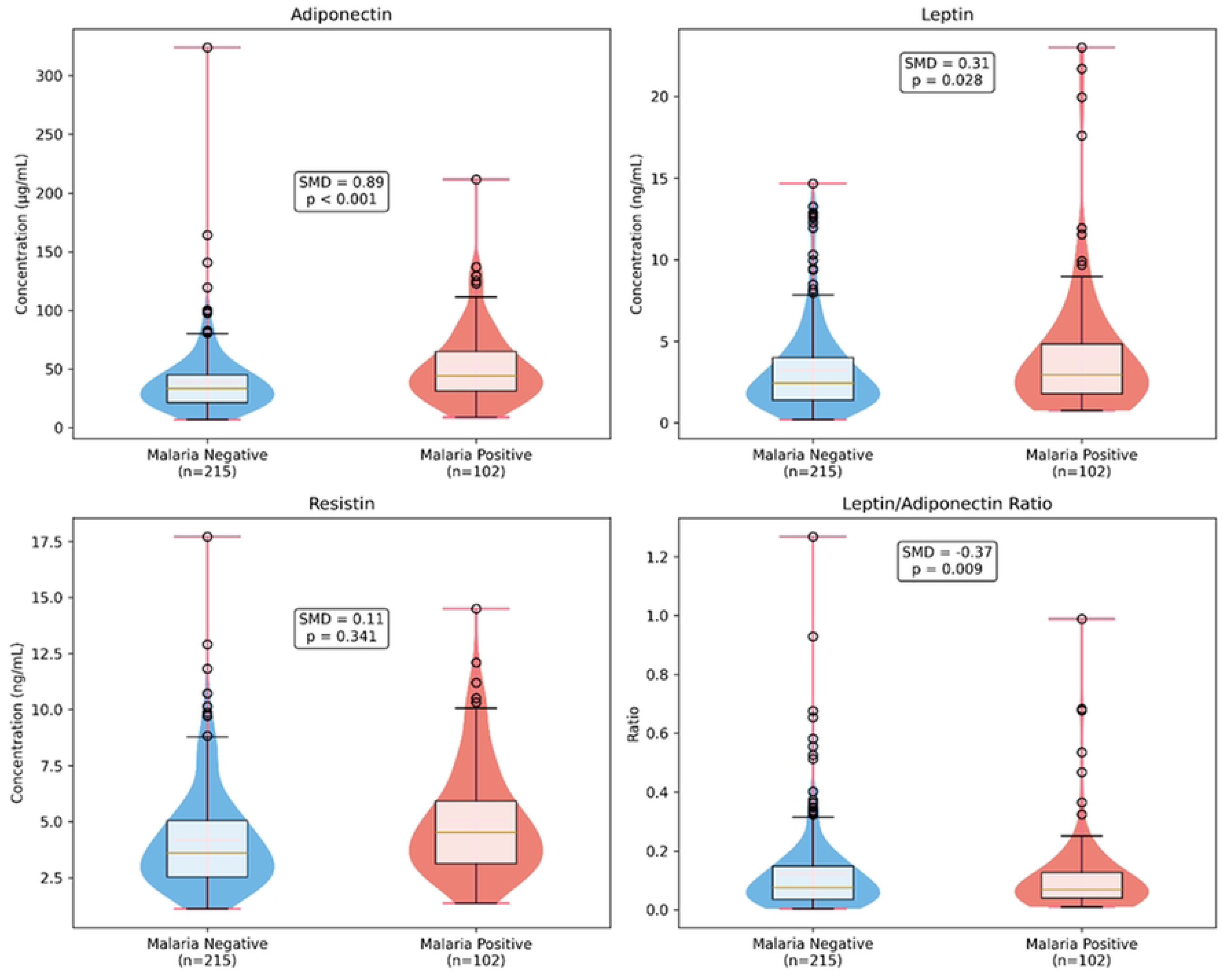
Adipocytokine distributions by malaria status

**Table 2:** Adipocytokine concentrations by malaria status.

| Adipocytokine | Overall<br>(n=317) | Malaria<br>Negative<br>(n=215) | Malaria<br>Positive<br>(n=102) | SMD<br>(95%<br>CI) | P-value | FDR-adjusted<br>P |
| --- | --- | --- | --- | --- | --- | --- |
| Adiponectin<br>($\mu\text{g/mL}$) | | | | | | |
| Geometric mean<br>(95% CI) | 36.8<br>(34.2-39.6) | 32.1 (29.4-35.1) | 45.2<br>(39.8-51.3) | 0.89<br>(0.61-1.17) | <0.001 | <0.001 |
| Median (IQR) | 38.2<br>(24.6-56.8) | 33.1 (22.4-48.9) | 47.8<br>(32.1-68.5) |  |  |  |
| Range | 8.9-156.3 | 8.9-142.1 | 15.2-156.3 |  |  |  |
| Leptin (ng/mL) |  |  |  |  |  |  |
| Geometric mean<br>(95% CI) | 2.5 (2.2-2.8) | 2.3 (2.0-2.6) | 2.8 (2.3-3.4) | 0.31<br>(0.03-0.59) | 0.028 | 0.084 |
| Median (IQR) | 2.4 (1.2-4.8) | 2.2 (1.1-4.2) | 2.9 (1.5-5.8) |  |  |  |
| Range | 0.2-18.7 | 0.2-16.4 | 0.4-18.7 |  |  |  |
| Resistin (ng/mL) |  |  |  |  |  |  |
| Geometric mean<br>(95% CI) | 3.9 (3.6-4.2) | 3.8 (3.5-4.2) | 4.1 (3.6-4.7) | 0.11 (-<br>0.17-0.39) | 0.341 | 0.511 |
| Median (IQR) | 3.8 (2.6-5.9) | 3.7 (2.5-5.7) | 4.0 (2.8-6.2) |  |  |  |
| Range | 0.8-15.2 | 0.8-15.2 | 1.1-12.8 |  |  |  |
| Leptin/Adiponectin<br>Ratio |  |  |  |  |  |  |
| Geometric mean<br>(95% CI) | 0.068<br>(0.062-0.075) | 0.072<br>(0.064-0.081) | 0.062<br>(0.051-0.075) | -0.37 (-<br>0.65 to<br>-0.09) | 0.009 | 0.027 |
| Median (IQR) | 0.066<br>(0.035-0.125) | 0.071<br>(0.038-0.135) | 0.058<br>(0.030-0.108) |  |  |  |
| Range | 0.005-0.892 | 0.005-0.892 | 0.008-0.456 |  |  |  |

*Geometric means (GM) and 95% CI computed on log-transformed values. Differences are GM ratios (Malaria-positive vs Malaria-negative); SMD is Cohen's d with 95% CI. P-values from Mann–Whitney U tests; FDR correction by Benjamini–Hochberg applied across the four primary adipocytokine comparisons.*

Leptin showed a modest unadjusted elevation (geometric mean 2.8 ng/mL [95% CI 2.3–3.4] vs 2.3 ng/mL [95% CI 2.0–2.6]; p=0.028), but this did not remain significant after Benjamini– Hochberg correction across the adipocytokine family (FDR-adjusted p=0.084). Resistin did not differ significantly between groups (4.1 vs 3.8 ng/mL; p=0.341).

### Correlations with parasite density (malaria-positive only)

Among malaria-positive children with non-zero parasite density, adiponectin increased with parasite density (Fig 2; Spearman ρ=0.31; 95% CI 0.12–0.47; p=0.002). This association remained significant after FDR correction within the parasite-density correlation family (FDR-adjusted p=0.008). Correlations for leptin, resistin, and L/A were smaller and did not survive FDR correction. Complete correlation results are shown in S2 Table.

**Fig 2.**
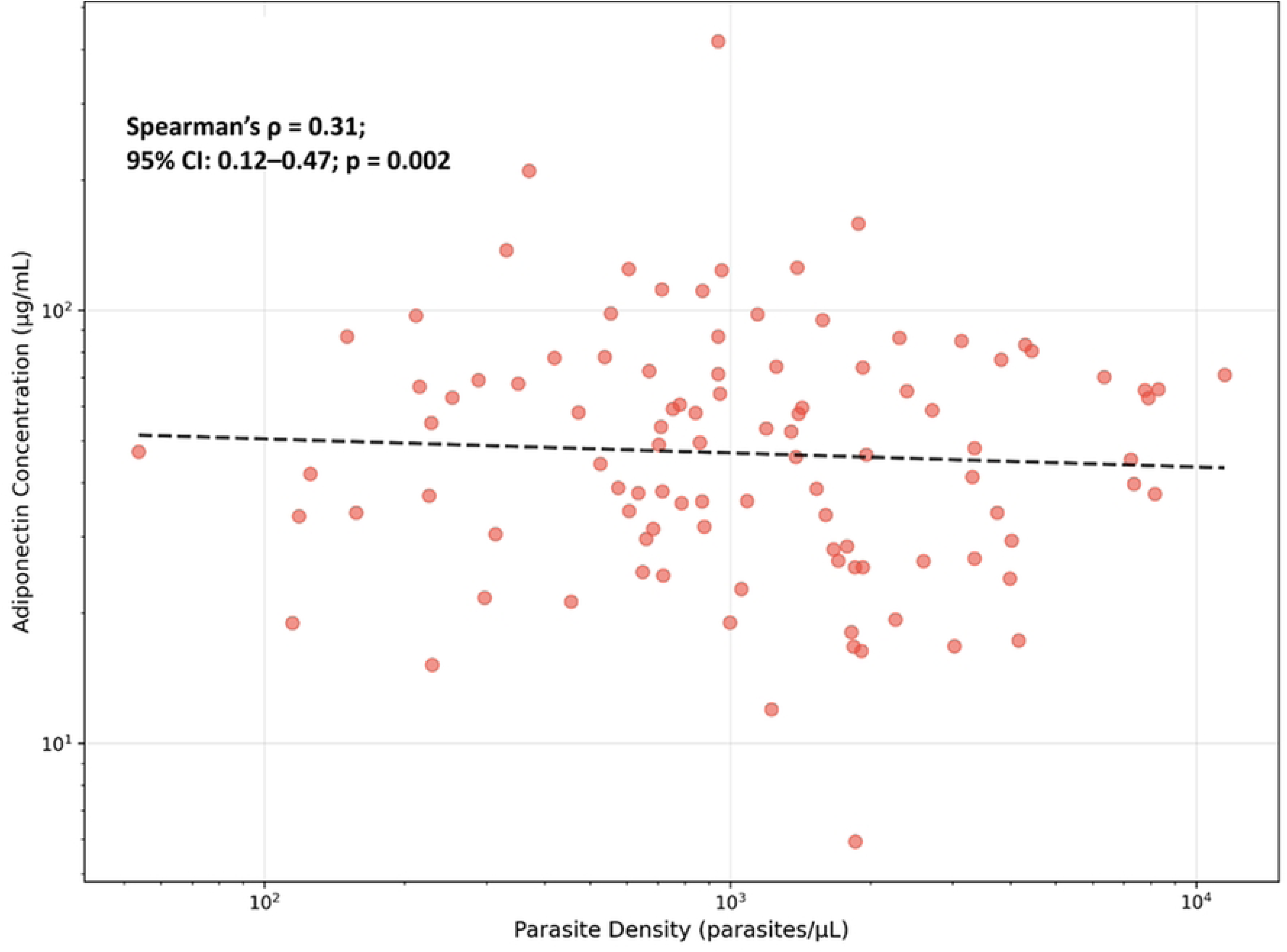
Adiponectin–parasite density relationship in asymptomatic malaria (positives only)

### Multivariable regression analysis

In multivariable linear models of log-adipocytokines with cluster-robust standard errors (cluster: school), adiponectin remained higher and L/A lower among malaria-positive children (Table 3). In the fully adjusted model (Model 3), malaria-positive status was associated with +42% adiponectin (95% CI: 18–71; *p*<0.001) and −31% L/A (95% CI: −49 to −7; *p* = 0.015).

**Table 3:**
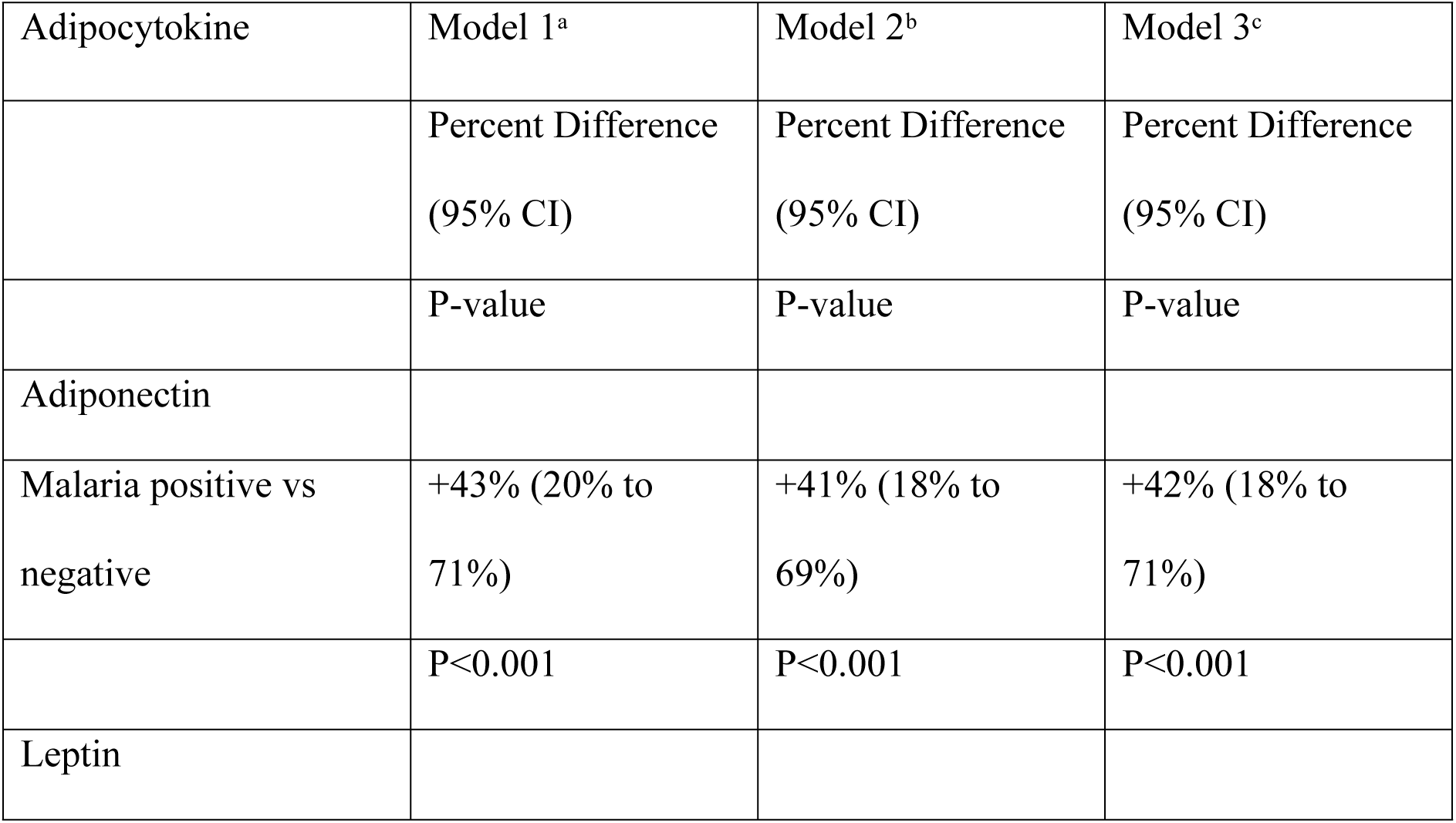
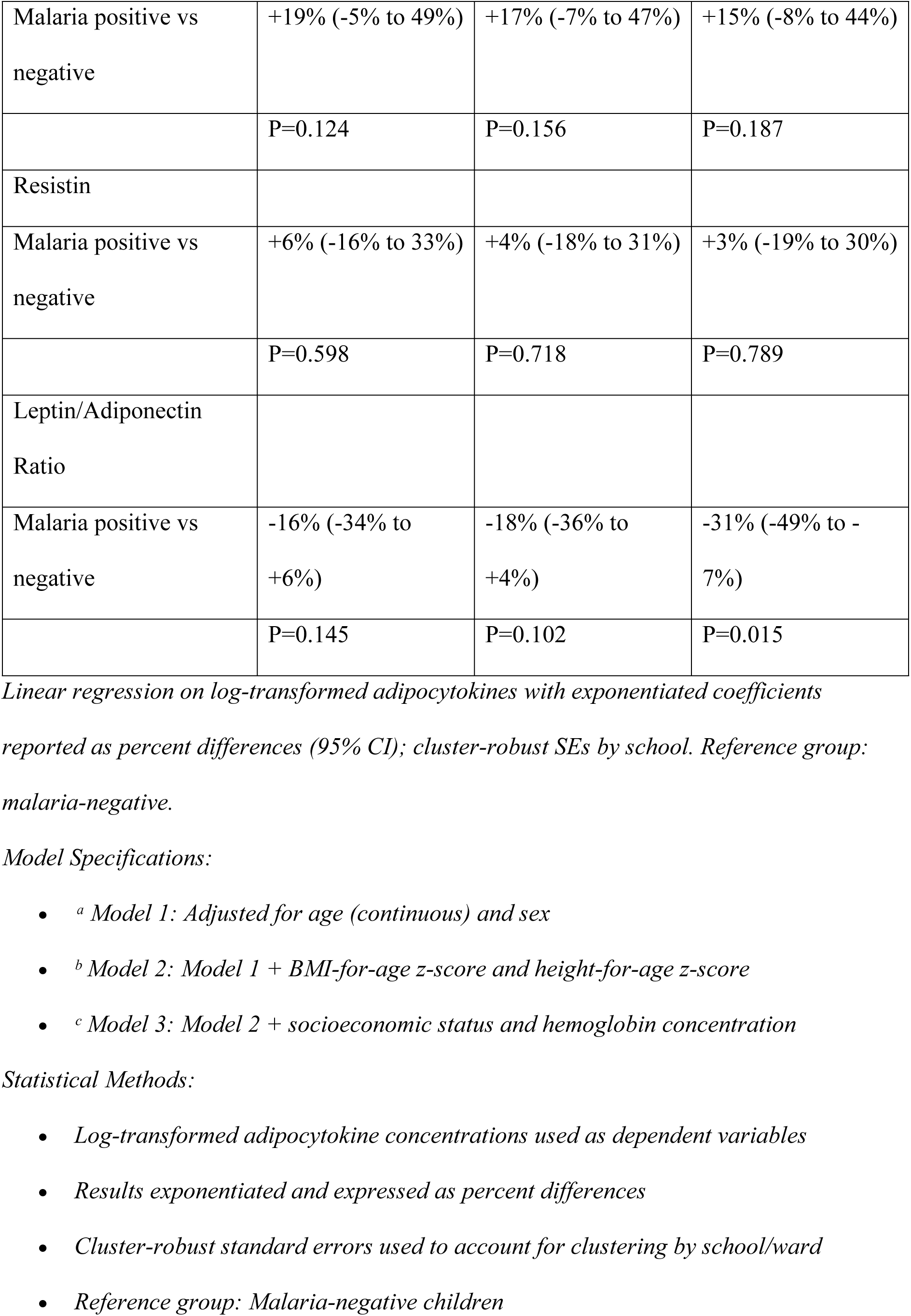
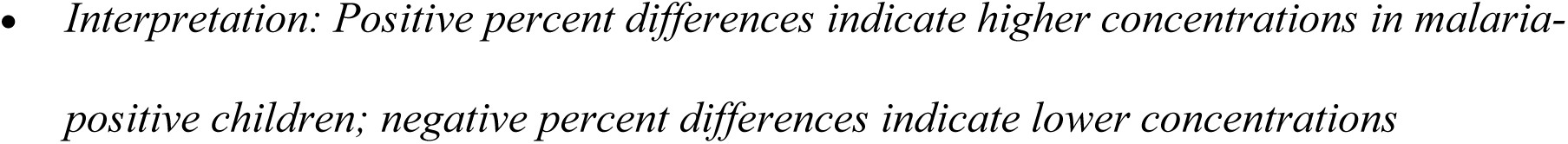
Multivariable Regression Models for Associations Between Malaria Status and Adipocytokine Concentrations.

| Adipocytokine | Model 1<sup>a</sup> | Model 2<sup>b</sup> | Model 3<sup>c</sup> |
| --- | --- | --- | --- |
|  | Percent Difference<br>(95% CI) | Percent Difference<br>(95% CI) | Percent Difference<br>(95% CI) |
|  | P-value | P-value | P-value |
| Adiponectin |  |  |  |
| Malaria positive vs<br>negative | +43% (20% to<br>71%) | +41% (18% to<br>69%) | +42% (18% to<br>71%) |
| | $P < 0.001$ | $P < 0.001$ | $P < 0.001$ |
| Leptin |  |  |  |
| Malaria positive vs negative | +19% (-5% to 49%) | +17% (-7% to 47%) | +15% (-8% to 44%) |
|  | P=0.124 | P=0.156 | P=0.187 |
| Resistin |  |  |  |
| Malaria positive vs negative | +6% (-16% to 33%) | +4% (-18% to 31%) | +3% (-19% to 30%) |
|  | P=0.598 | P=0.718 | P=0.789 |
| Leptin/Adiponectin Ratio |  |  |  |
| Malaria positive vs negative | -16% (-34% to +6%) | -18% (-36% to +4%) | -31% (-49% to -7%) |
|  | P=0.145 | P=0.102 | P=0.015 |

*Linear regression on log-transformed adipocytokines with exponentiated coefficients* *reported as percent differences (95% CI); cluster-robust SEs by school. Reference group:* *malaria-negative.*
*Model Specifications:*
• <sup>a</sup> *Model 1: Adjusted for age (continuous) and sex* • <sup>b</sup> *Model 2: Model 1 + BMI-for-age z-score and height-for-age z-score* • <sup>c</sup> *Model 3: Model 2 + socioeconomic status and hemoglobin concentration*
*Statistical Methods:*
• *Log-transformed adipocytokine concentrations used as dependent variables* • *Results exponentiated and expressed as percent differences* • *Cluster-robust standard errors used to account for clustering by school/ward* • *Reference group: Malaria-negative children*
• *Interpretation: Positive percent differences indicate higher concentrations in malaria-* *positive children; negative percent differences indicate lower concentrations*

Leptin and resistin were not independently associated with malaria status after adjustment (all adjusted *p* > 0.05); for leptin, the adjusted percent difference was +15% (95% CI: −8 to 44; *p* = 0.187). Report Model 1–3 as shown in Table 3 and emphasize adiponectin↑ and L/A↓ as the robust findings.

### Multivariable regression analysis

In multivariable linear models of log-adipocytokines with cluster-robust standard errors (cluster: school), adiponectin remained higher and L/A lower among malaria-positive children (Table 3). In the fully adjusted model (Model 3), malaria-positive status was associated with +42% adiponectin (95% CI 18–71; p<0.001) and −31% L/A (95% CI −49 to −7; p=0.015).

### Correlation matrix (overall)

Pairwise Spearman correlations among adipocytokines, anthropometry, and hematologic indices are summarized in Table 4. FDR-significant pairs are flagged in the table; other correlations should be interpreted as exploratory.

**Table 4.**
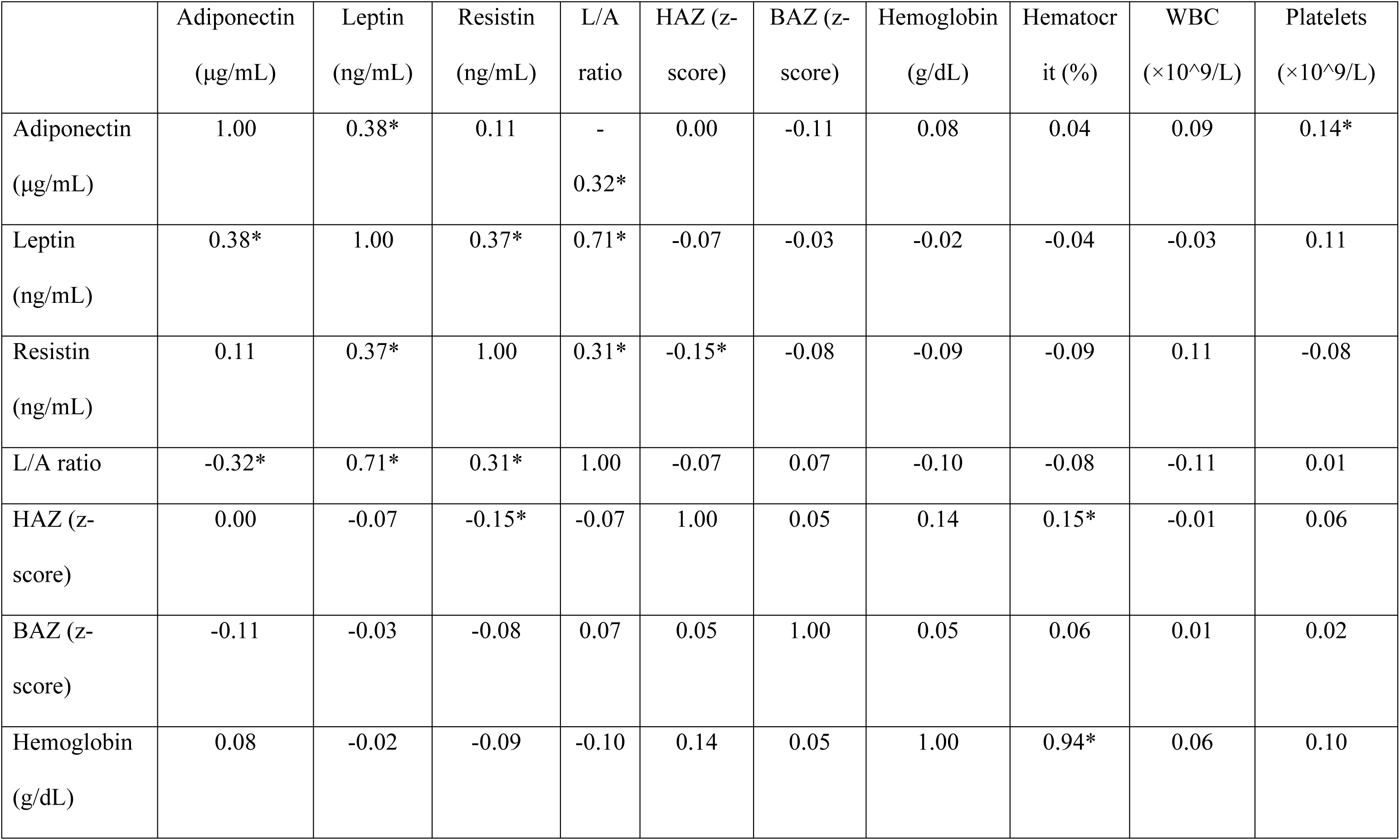
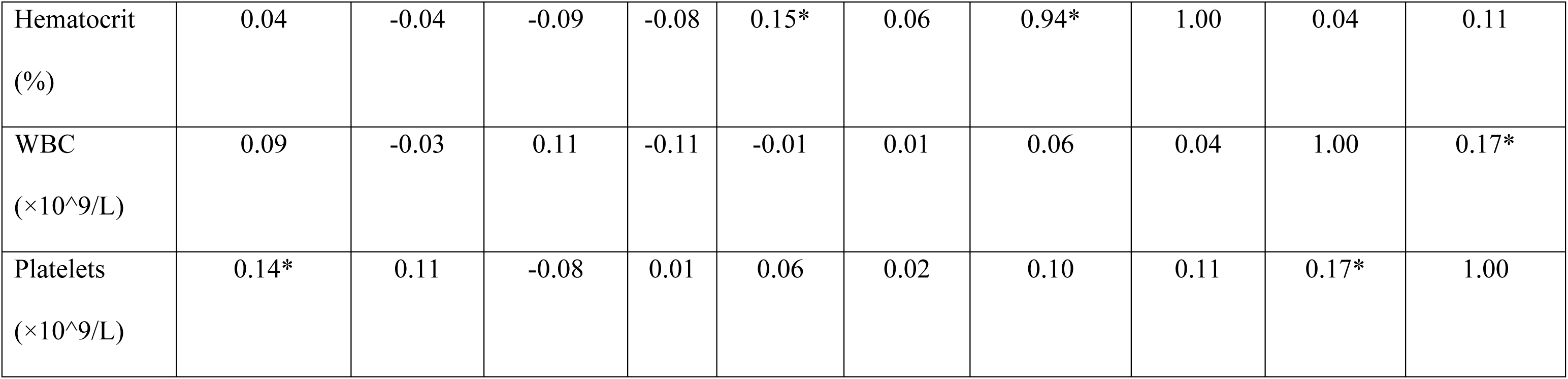
Spearman correlation matrix (FDR-significant pairs starred)

| | Adiponectin<br>($\mu\text{g/mL}$) | Leptin<br>($\text{ng/mL}$) | Resistin<br>($\text{ng/mL}$) | L/A<br>ratio | HAZ (z-score) | BAZ (z-score) | Hemoglobin<br>($\text{g/dL}$) | Hematocr<br>it (%) | WBC<br>($\times 10^9/\text{L}$) | Platelets<br>($\times 10^9/\text{L}$) |
| --- | --- | --- | --- | --- | --- | --- | --- | --- | --- | --- |
| Adiponectin<br>($\mu\text{g/mL}$) | 1.00 | 0.38* | 0.11 | -<br>0.32* | 0.00 | -0.11 | 0.08 | 0.04 | 0.09 | 0.14* |
| Leptin<br>($\text{ng/mL}$) | 0.38* | 1.00 | 0.37* | 0.71* | -0.07 | -0.03 | -0.02 | -0.04 | -0.03 | 0.11 |
| Resistin<br>($\text{ng/mL}$) | 0.11 | 0.37* | 1.00 | 0.31* | -0.15* | -0.08 | -0.09 | -0.09 | 0.11 | -0.08 |
| L/A ratio | -0.32* | 0.71* | 0.31* | 1.00 | -0.07 | 0.07 | -0.10 | -0.08 | -0.11 | 0.01 |
| HAZ (z-score) | 0.00 | -0.07 | -0.15* | -0.07 | 1.00 | 0.05 | 0.14 | 0.15* | -0.01 | 0.06 |
| BAZ (z-score) | -0.11 | -0.03 | -0.08 | 0.07 | 0.05 | 1.00 | 0.05 | 0.06 | 0.01 | 0.02 |
| Hemoglobin<br>($\text{g/dL}$) | 0.08 | -0.02 | -0.09 | -0.10 | 0.14 | 0.05 | 1.00 | 0.94* | 0.06 | 0.10 |
| Hematocrit<br>(%) | 0.04 | -0.04 | -0.09 | -0.08 | 0.15* | 0.06 | 0.94* | 1.00 | 0.04 | 0.11 |
| WBC<br>($\times 10^9/L$) | 0.09 | -0.03 | 0.11 | -0.11 | -0.01 | 0.01 | 0.06 | 0.04 | 1.00 | 0.17* |
| Platelets<br>($\times 10^9/L$) | 0.14* | 0.11 | -0.08 | 0.01 | 0.06 | 0.02 | 0.10 | 0.11 | 0.17* | 1.00 |

### Subgroup and sensitivity analyses

Age-stratified analyses suggested a larger adiponectin difference in older children (≥5 years) (interaction p=0.032), with a weaker, non-significant association in younger children (<5 years). Patterns were similar by sex (interaction p=0.671); visual summaries are provided in S3 Fig. Findings were robust in CRP-restricted analyses (CRP ≤3 mg/L), supporting specificity to subclinical infection rather than acute inflammation (S4 Fig). Results restricted to microscopy-confirmed malaria were concordant with the primary analyses: adiponectin remained elevated and the adiponectin–parasite density correlation persisted (ρ≈0.31; p=0.006).

## Discussion

### Principal findings

In this community-based sample of Nigerian schoolchildren, asymptomatic malaria was associated with a distinct metabolic signature characterized by higher adiponectin and a lower leptin-to-adiponectin (L/A) ratio, patterns that persisted after covariate adjustment. In fully adjusted models, malaria-positive status corresponded to +42% adiponectin (95% CI: 18–71; p<0.001) and −31% L/A (95% CI: −49 to −7; p = 0.015). By contrast, leptin and resistin were not independently associated with malaria after adjustment, and the modest unadjusted elevation in leptin did not survive false-discovery-rate correction. Among malaria-positive children, adiponectin increased with parasite density, underscoring that “silent” infections are metabolically active.

### Interpretation in the framework of immunometabolism

A converging literature shows that chronic or recurrent immune activation reshapes endocrine control of energy balance, immunometabolism, and, reciprocally, adipose-derived hormones modulate immune tone [7]. Within this interface, adipocytokines operate as integrators of metabolism and inflammation [8]. Leptin regulates appetite and supports pro-inflammatory responses [9,10], adiponectin enhances insulin sensitivity and generally exerts anti-inflammatory actions [11,12], and resistin participates in inflammatory/insulin-resistance pathways [13]. Our pattern, adiponectin↑ with L/A↓, is therefore consistent with a systemic counter-inflammatory/metabolic-flexibility response to persistent low-grade antigenic stimulation from subclinical parasitemia [1–6,7–8,11–12]. The dose-response between parasite density and adiponectin further supports a graded physiological adaptation rather than a binary effect.

### Clarifying the role of leptin

Although leptin was modestly higher in unadjusted comparisons, it was not significant after FDR correction and did not remain significant in multivariable models. In the asymptomatic pediatric state, leptin thus does not show an independent association with malaria. This contrasts with patterns sometimes reported in acute, symptomatic disease and adult cohorts, contexts where acute-phase signaling may dominate, and highlights the importance of disease phase and age in interpreting adipocytokines in malaria biology [2,6,8–12]. Focusing inference on adiponectin and L/A avoids over-attribution to leptin and reflects the most robust signals in our data.

### Age patterning

The stronger adiponectin difference in older children (≥5 y) is biologically plausible. Across middle childhood, adipose tissue and endocrine-immune crosstalk continue to mature, potentially permitting a larger adiponectin response to sustained parasitemia [7–9,11–12]. Older children in endemic settings also accumulate exposure, which could amplify immunometabolic adaptation. These hypotheses warrant longitudinal testing.

### Public health and clinical implications

First, the findings strengthen the view that asymptomatic malaria is not physiologically benign [1–6]. The combination of adiponectin↑, L/A↓, and a density-dependent gradient implies a hidden metabolic burden that may interact with nutrition and anaemia in school-age children. Second, adiponectin (and derived L/A) may serve as responsive, non-invasive indicators of subclinical infection intensity or resolution, subject to validation of specificity and within-child dynamics [7–8,11–12]. Third, these results support integrated strategies that couple malaria control with child metabolic health, complementing policies and arguments recognizing the public-health relevance of asymptomatic infections [1–4].

### Strengths and limitations

Strengths include community-based sampling, prespecified models with cluster-robust SEs (school as cluster), duplicate ELISAs with quality control, and sensitivity analyses (microscopy restriction; CRP ≤3 mg/L), in line with STROBE reporting (Methods). Limitations warrant caution. The cross-sectional design precludes causal inference; longitudinal follow-up around infection and treatment is needed. Although we excluded overt illness and performed a CRP-restricted analysis, residual low-grade inflammation from other causes cannot be excluded. We did not include direct metabolic endpoints (e.g., glucose/insulin dynamics), which would sharpen mechanism. Finally, results from a single LGA in southwest Nigeria require evaluation in other epidemiologic contexts; nonetheless, the internal consistency across Tables 2–3, Fig 2, and Supplement argue for robustness.

### Future directions

Priority studies include: (i) longitudinal designs to define temporal coupling between parasite dynamics and adipocytokines; (ii) metabolic phenotyping (glucose–insulin homeostasis, lipid flux) to test whether adiponectin-dominant signaling is adaptive or entails trade-offs; (iii) intervention studies to determine whether treating asymptomatic malaria normalizes adipocytokines and improves clinical outcomes; and (iv) implementation research integrating metabolic monitoring into school-based malaria control where cost-effective. At a policy level, these data support evaluating school-based screen-and-treat approaches for asymptomatic malaria, optionally coupled with simple metabolic surveillance, particularly in older children, with implementation tailored to local epidemiology and resources

## Conclusions

Asymptomatic malaria in children is accompanied by elevated adiponectin and reduced L/A, with adiponectin tracking parasite density. Our findings support the view that subclinical malaria is metabolically active in children. The null adjusted results for leptin and resistin refine a parsimonious, reproducible signal, adiponectin↑/L-A↓, for future validation and translation [7–12].

## Data Availability

All data underlying the findings of this study, comprising the minimally sufficient de-identified dataset, analysis code, and a detailed data dictionary, are publicly available on Zenodo at DOI: 10.5281/zenodo.17162616. No special permissions are required to access these materials, and no potentially identifying information is included.

## Acknowledgments

We thank the children and families who participated in this study, as well as the community leaders and local health workers who facilitated recruitment and data collection. We acknowledge the dedicated work of our field teams and laboratory personnel. We are grateful to the Oyo State Ministry of Health for their support and collaboration. All individuals named in the acknowledgments provided permission to be acknowledged.

## Author contributions (CRediT)

AEO: Conceptualization; Methodology; Supervision; Writing – original draft; Visualization; Writing-review & editing. OJA: Data curation; Investigation; Writing-review & editing. KIA: Data curation; Investigation; Writing-review & editing. BEO: Conceptualization; Formal analysis; Software; Visualization; Writing-original draft; Writing – review & editing.

## Supporting information

S1 Checklist. STROBE checklist for cross-sectional studies.

S1 Table. Baseline characteristics by malaria status (per-variable denominators, tests, and effect sizes).

S2 Table. Spearman correlation coefficients among adipocytokines, anthropometry, and hematologic indices (FDR-adjusted p-values indicated).

S3 Table. Multivariable model specifications and covariate definitions (Models 1–3).

---

**S1 Fig.**
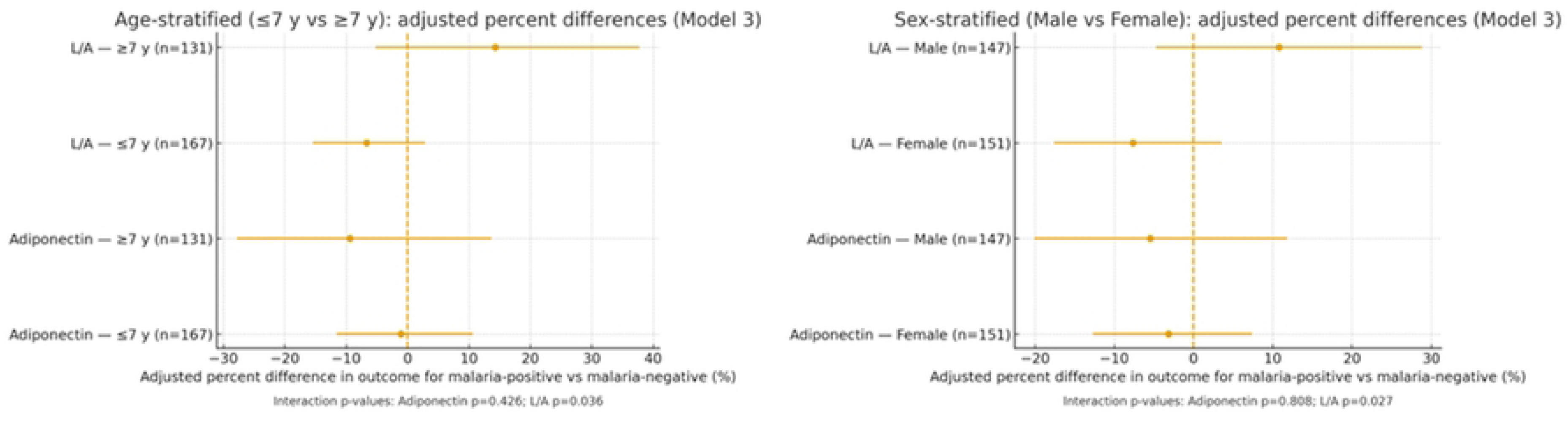
Age- and sex-stratified associations of asymptomatic malaria with adiponectin and leptin-to-adiponectin ratio (Model 3; cluster-robust SEs).

**S2 Fig.**
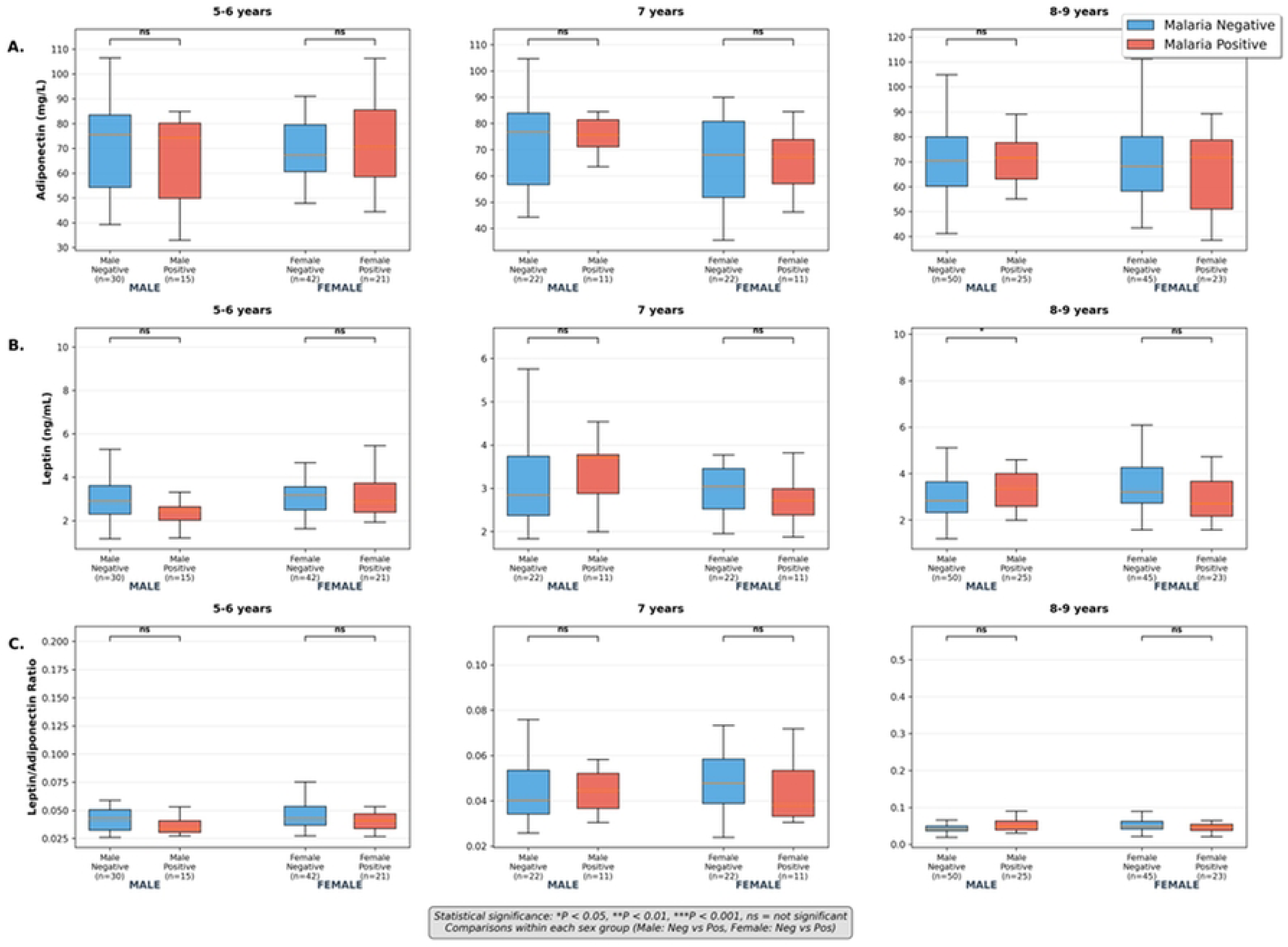
Age-stratified boxplots of adiponectin, leptin, and leptin-to-adiponectin ratio by malaria status.

**S3 Fig.**
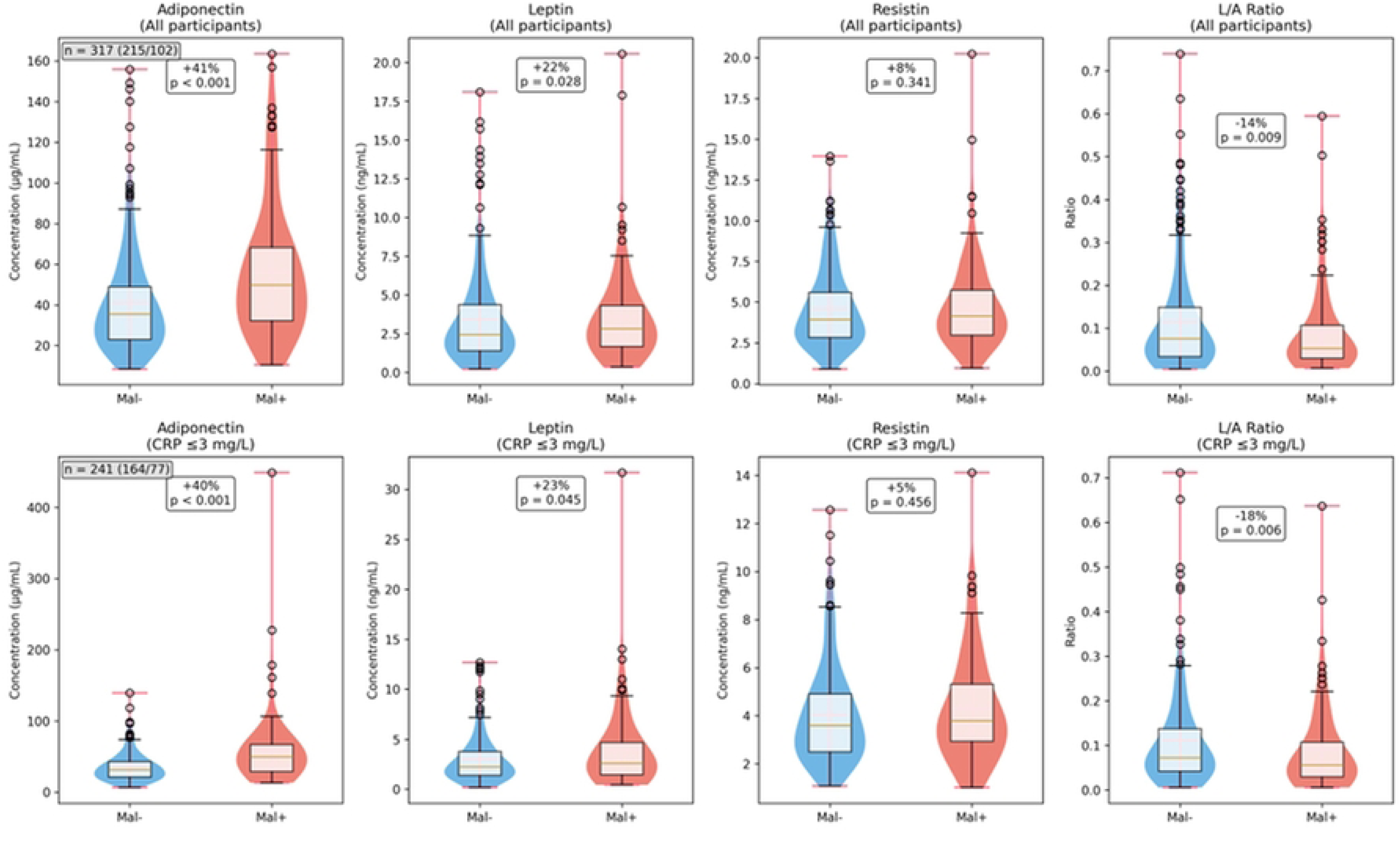
Primary outcomes restricted to CRP ≤3 mg/L subsample (violin/box overlays with percent differences and p-values).

